# Behavioral changes before lockdown, and decreased retail and recreation mobility during lockdown, contributed most to the successful control of the COVID-19 epidemic in 35 Western countries

**DOI:** 10.1101/2020.06.20.20136382

**Authors:** Koen Deforche, Jurgen Vercauteren, Viktor Müller, Anne-Mieke Vandamme

## Abstract

The COVID-19 pandemic has prompted a lockdown in many countries to control the exponential spread of the SARS-CoV-2 virus (*1, 2*). This resulted in curbing the epidemic by reducing the time-varying basic reproduction number (*R*_*t*_) to below one (*3, 4*). Governments are looking for evidence to balance the demand of their citizens to ease some of the restriction, against the fear of a second peak in infections. More details on the specific circumstances that promote exponential spread (i.e. *R*_*t*_ *>* 1) and the measures that contributed most to a reduction in *R*_*t*_ are needed. Here we show that in 33 of 35 Western countries (32 European, plus Israel, USA and Canada), *R*_*t*_ fell to around or below one during lockdown (March – May 2020). One third of the effect happened already on average 6 days before the lockdown, with lockdown itself causing another major drop in transmission. Country-wide compulsory usage of masks was implemented only in Slovakia 10 days into lockdown, and on its own reduced transmission by half. During lockdown, decreased mobility in retail and recreation was an independent predictor of lower *R*_*t*_ during lockdown, while changes in other types of mobility were not. These results are consistent with anecdotal evidence that large recreational gatherings are super-spreading events (*5, 6*), and may even suggest that infections during day-to-day contact at work are not sufficient to spark exponential growth. Our data suggest measures that will contribute to avoiding a second peak include a tight control on circumstances that facilitate massive spread such as large gatherings especially indoors, physical distancing, and mask use.

---

In February-March 2020, it became clear that the COVID-19 pandemic had been introduced in Europe and Northern America (*7*) and governments and societies have responded in various ways to slow down the spread of the virus. In the containment phase, testing, contact-tracing and individual quarantine of (suspected) infected individuals are crucial to prevent a widespread epidemic. If this fails, mitigation measures consist mainly of mobility restrictions and encouraged or mandated physical distancing to everyone regardless of symptoms or testing results (jointly called lockdown).

Failing containment, most European countries managed to curb the epidemic by May through lockdown measures (timeline and details of measures in the different countries (*1*)), however without fully understanding which of the different measures had the most impact. Because of economic and social implications of a lockdown, governments are cautiously relaxing measures while trying to avoid a second peak by closely following the effect on the epidemics through testing and monitoring hospitalizations and casualties (*8*). Coming out of lockdown entails a return to containment measures, with testing, contact tracing and quarantine again being crucial.

Epidemiological models have been widely used to make predictions about the evolution of the COVID-19 epidemics, to predict the outcome of testing strategies and intervention scenarios, and to estimate epidemiological parameters (*9–13*). Epidemic spread is determined by both biological properties of the virus as well as the behavior of the host population, and this is reflected in the epidemiological parameters. One important parameter, the time-varying basic reproductive number *R*_*t*_, determines whether an epidemic is exponentially growing (*R*_*t*_ *>* 1) or declining (*R*_*t*_ *<* 1). The goal of both containment and mitigation measures is to bring *R*_*t*_ below 1 by changing the behavior of the infected, respectively, susceptible population through reducing their contact networks.

In Western countries the population was developing an increased awareness as soon as media reported on the epidemic unfolding in Lombardy (Italy), and this led to a gradual response ranging from voluntary physical distancing to government imposed mandatory self-isolation upon showing symptoms, banning of public events, and encouraging physical distancing, with ultimately a drastic lockdown when many businesses and schools were closed and mobility was severely restricted (*14*). The diversity of measures and their timing gives us the opportunity to investigate which differences were correlated with a better outcome. However, reliable data quantifying the impact on human behavior of various country-dependent specific lockdown measures is currently lacking.

Attempts to create such data sets face challenges such as variation in compliance to imposed measures, and the unknown level of measures taken by individuals or organizations in absence or anticipation of government regulations. Rather than an account of the ordered measures in each country, we used mobility changes extracted from smartphone location data as an objective indication and quantification of the lockdown (Suppl 1).

From incidence data of deaths and diagnosed cases in 35 countries (ECDC data downloaded on 3 June 2020, see Suppl 2), we estimated the change in transmission over time, while differentiating between the effect of measures that preceded the lockdown, and the effect of the lockdown itself. Since incidence data of deaths is considered more reliable than of diagnosed cases (which is also highly dependent on testing policy and test availability), we estimated suitable values for the dispersion parameters by estimating variance from the fits and then use a slightly more cautious value (Suppl 3). For case data we then simply decreased the value to give a higher weight to death data, favoring better fits of death data over case data. Only data up to 60 days into the lockdown (up to about mid-May in European countries) were used to avoid the confounding effect of the gradual lifting of restrictions, which started in many European countries in May. Given that time between infection and reported death was estimated as 24 (95% IQR 18 - 32) days, potential lifting of measures in the beginning of May for some countries could not have confounded the estimates.

In order to obtain comparable estimates of *R*_*t*_, a simple SEIR compartment model was used, with most epidemiological parameters kept constant. Parameters that model transmission rates were allowed to change from an initial estimated value *R*_*t*,0_ during a transition period also estimated from the data, to *R*_*t*,1_ until the day that mobility changes started, and then to *R*_*t*,2_ during the lockdown identified on the basis of mobility data, using a piecewise linear model (Figure 1a and methods). The estimated models fitted well the reported incidence data for most countries (see Suppl 4). Across all countries, the median of posterior estimates for *R*_*t*,0_ was 3.6 (95% IQR 2.5 – 4.9) (see Figure 2a). Before changes in mobility were observed (*d*_1_), the reproduction number was reduced to 2.3 (95% IQR 1.7 – 3.0). During lockdown, transmission was further reduced to 0.77 (95% IQR 0.58 – 1.04). Only for Belarus and Moldova, the estimate of *R*_*t*,2_ was higher than 1 (*>* 95% credible), while all other countries have the *R*_*t*,2_ value below or around 1. Estimates for *R*_*t*,0_ are expected to vary depending on setting, methodology and assumptions on parameters (especially duration of infectious period and generation time) and assumptions on how the number may vary over time (*15*). We find that our estimated values for *R*_*t*,0_ tend to be higher, and estimated values for *R*_*t*,1_ lower, compared to estimates in other studies for *R*_0_ of around 2.7. Our estimates for *R*_*t*,0_ are similar to *R*_0_ estimates obtained using models that also consider interventions that preceded a full lockdown (*14*).

**Figure 1:**
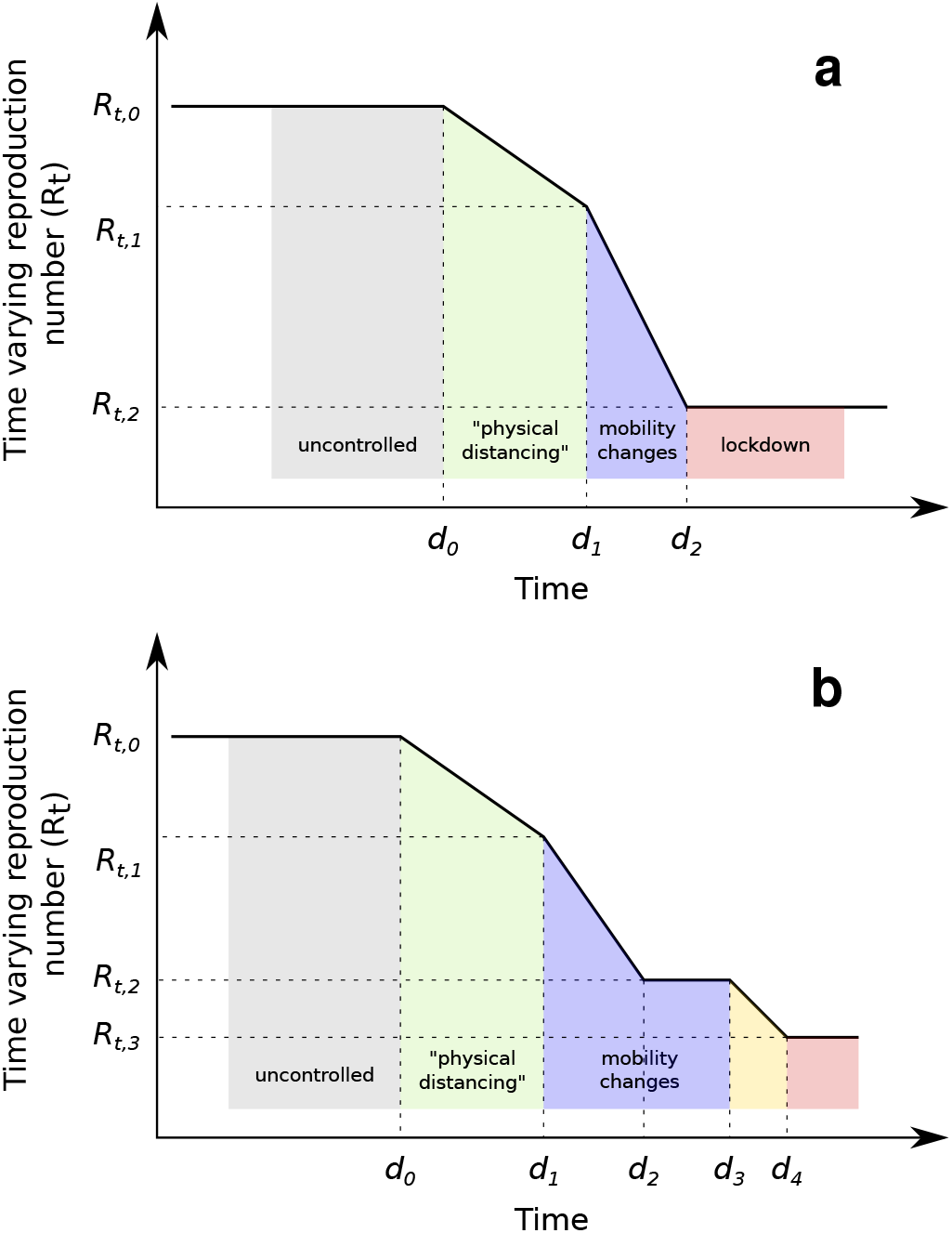
Model for changes of the time-varying reproduction number *R*_*t*_ as a piece-wise linear function. Dates *d*_1_ and *d*_2_ were estimated from mobility data. Date *d*_0_ and values for *R*_*t*,0_ – *R*_*t*,2_ were estimated from incidence data on diagnosed cases and deaths. a. Model used for all countries; b. Model additionally used for Slovakia, which allowed an extra change during lockdown with dates *d*_0_, *d*_3_ and *d*_4_, and values for *R*_*t*,0_ – *R*_*t*,3_ estimated from incidence data on diagnosed cases and deaths.

**Figure 2:**
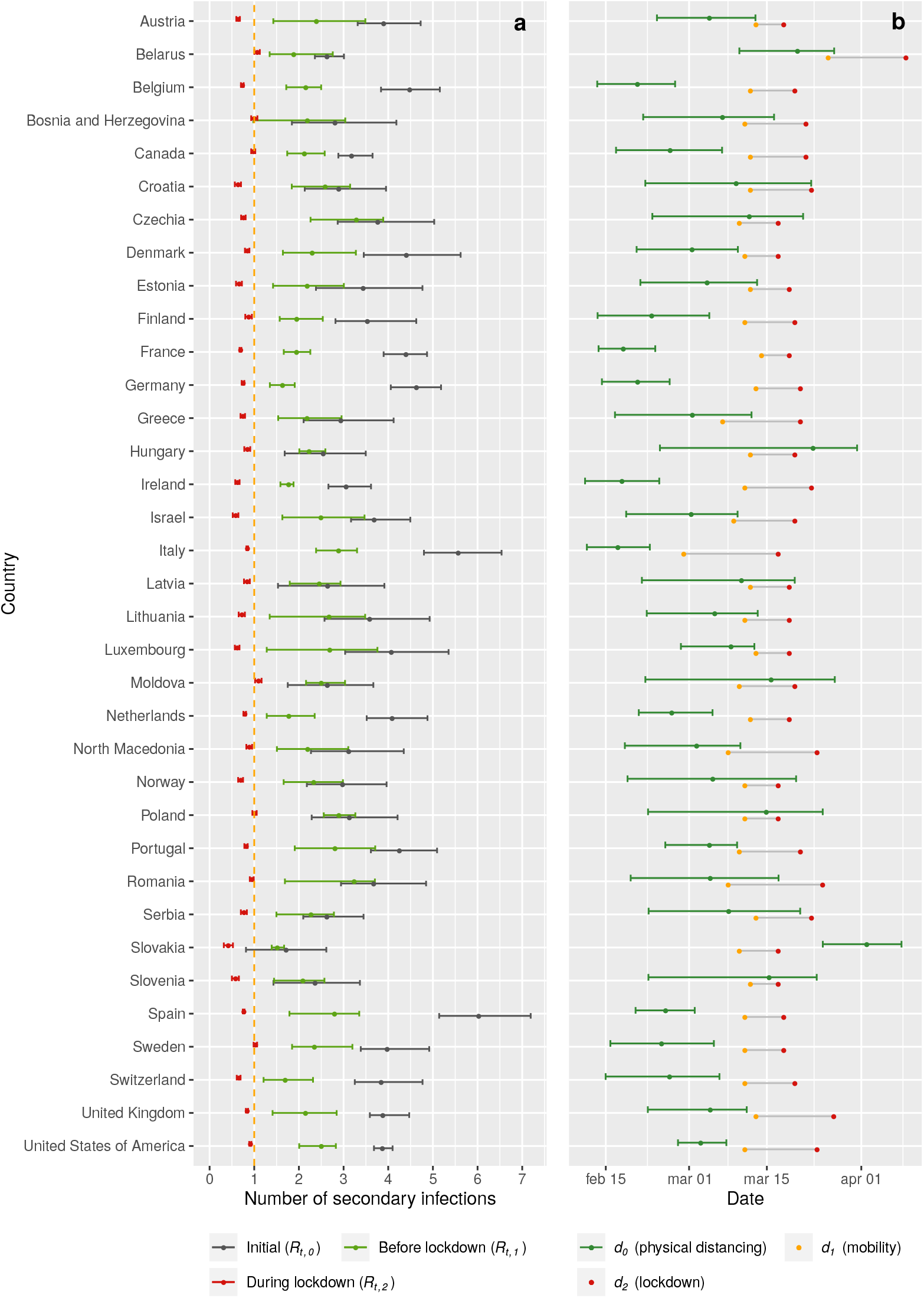
a. Posterior estimates of the initial basic reproduction numbers (*R*_*t*,0_), the reproduction number at start of lockdown (*R*_*t*,1_) and during lockdown (*R*_*t*,2_). b. Estimated median values (95% IQR) for *d*_0_ (date of first reduction in transmission, presumably due to physical distancing, estimated from incidence data of deaths and diagnosed cases); and lockdown transition start and end dates *d*_1_ and *d*_2_, estimated from mobility data (see also Suppl 1).

We estimated that the date *d*_0_ of the first decline of transmission preceded observed mobility changes (*d*_1_) with on average 6 days (95% IQR 0 – 22) (Figure 2b). During this period less invasive measures such as the mandatory self-isolation upon showing symptoms, banning of public events, and encouraging physical distancing may have contributed to a decline in transmission (there was no substantial decline in mobility during this period). The earliest of these dates (for Italy, Ireland, Germany, France, and Belgium) were all estimated around 20 February (95% IQR 12 – 25 February), which might have been a result of awareness raised by the discovery of the first European cluster around that time in Lombardy, Italy. Although the semantic meaning given to *d*_0_ (first date of decline of *R*_0_, before mobility changes), assumed that it would be estimated before *d*_1_, this order was not enforced. For Slovakia, the estimated model placed this date around 1 April (95% IQR March 22 – April 7), later than the dates *d*_1_ and *d*_2_ that mark the mobility changes period (10 – 17 March). At the same time, a suspiciously low value of 1.7 (95% IQR 0.8 – 2.6) for *R*_*t*,0_ was estimated. Both observations indicated that the assumption that mobility changes leading to the lockdown were the final measure to control transmission, was not applicable to data of Slovakia. We therefore re-estimated a model which allowed a further reduction in transmission to a value *R*_*t*,3_ (assuming a prior distribution *R*_*t*,3_ *− R*_*t*,2_ = *N* (0, 1) expressing no change with respect to *R*_*t*,2_), with a linear transition between co-estimated dates *d*_3_ and *d*_4_ (see Figure 1b). As prior distributions for *d*_0_, *R*_*t*,0_ –*R*_*t*,2_, and infection-to-death duration *µ*_*d*_, overall estimates obtained across all countries were used. This resulted in an additional estimated transition period for reduction in *R*_*t*_ between *d*_3_ of 30 March (95% IQR 18 March – 14 April), and *d*_4_ of 4 April (95% IQR 21 March – 18 April, Figure 3). This followed immediately after the introduction of mandatory face masks use in Slovakia on 25 March (*16*). This would imply that using face masks brought *R*_*t*_ from 0.87 (95% IQR 0.56 – 1.26) to 0.50 (95% IQR 0.30 – 0.70), or thus a 43% (95% IQR 0 – 64%) reduction in transmission. In further statistical analyses, the parameters estimated for this latter model were used for Slovakia.

**Figure 3:**
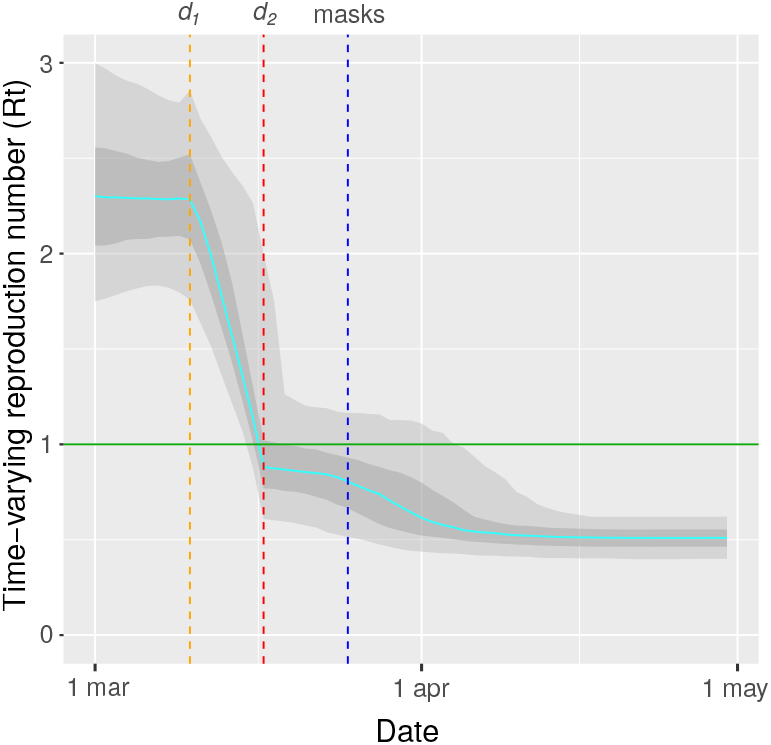
Estimated time-varying basic reproduction number *R*_*t*_ for Slovakia using a model that allowed an additional reduction of transmission rate at a co-estimated date during lockdown. The orange and red lines mark *d*_1_ and *d*_2_ as estimated from google mobility data. For reference, the date at which mandatory mask wearing was introduced (25 March) is indicated in blue.

To investigate how the reproduction number during lockdown *R*_*t*,2_ related to mobility changes, Google Mobility report data was used. For all six mobility categories, mobility during lock-down was significantly different compared to baseline (Wilcoxon paired test *p <* 0.01, data not shown). Lower *R*_*t*,2_ values during lockdown were significantly associated with a larger mobility reduction related to retail, recreation, and workplaces, and a lower mobility reduction related to residential (Figure 4), Table 1). The association of mobility reduction related to retail and recreation remained significant in a multivariate model which explained 40% of variance (adjusted *R*^2^) of the *R*_*t*,2_ value during lockdown (*p <* 0.002). This suggests that, in 35 Western countries, reductions of mobility related to retail and recreation during lockdown caused a mean reduction of *R*_*t*_ of 0.50 (95% CI 0.18 – 0.81), or thus an average reduction of 22% (95% CI 8 – 35) in transmission. Increased mobility related to parks may be an indicator of social restrictions in visiting friends or family, and explained up to 4% (95% CI 0 – 8) of additional reduction in transmission. The remaining reduction of *R*_*t*_ during lockdown may still be related to mobility changes of other types of mobility currently not reported by Google (for example mobility related to schools) or would require further refinement of categories to become observable. Alternatively, other types of changes that coincided in time with these mobility changes could be responsible.

**Table 1:** Univariate and multivariate associations of mobility changes during lockdown (per 10% mobility change) with the basic reproduction number *R*_*t*,2_ during lockdown. Mobility data related to transit stations and residential places were left out from the multivariate analysis since these variables were highly correlated with mobility data related to workplaces.

| Variable | Univariate estimate | Multivariate estimate |
| --- | --- | --- |
| Retail and recreation | 0.04 +/- 0.01 ($p < 0.004$) | 0.07 +/- 0.02 ($p < 0.004$) |
| Grocery and pharmacy | 0.02 +/- 0.02 | -0.04 +/- 0.03 |
| Parks | 0.004 +/- 0.006 | -0.018 +/- 0.009 ($p < 0.05$) |
| Transit stations | 0.03 +/- 0.02 |  |
| Workplaces | 0.06 +/- 0.02 ($p < 0.008$) | 0.04 +/- 0.03 |
| Residential | -0.11 +/- 0.04 ($p < 0.005$) | |
| $R_{t,1}$ | 0.05 +/- 0.06 | 0.02 +/- 0.05 |

**Figure 4:**
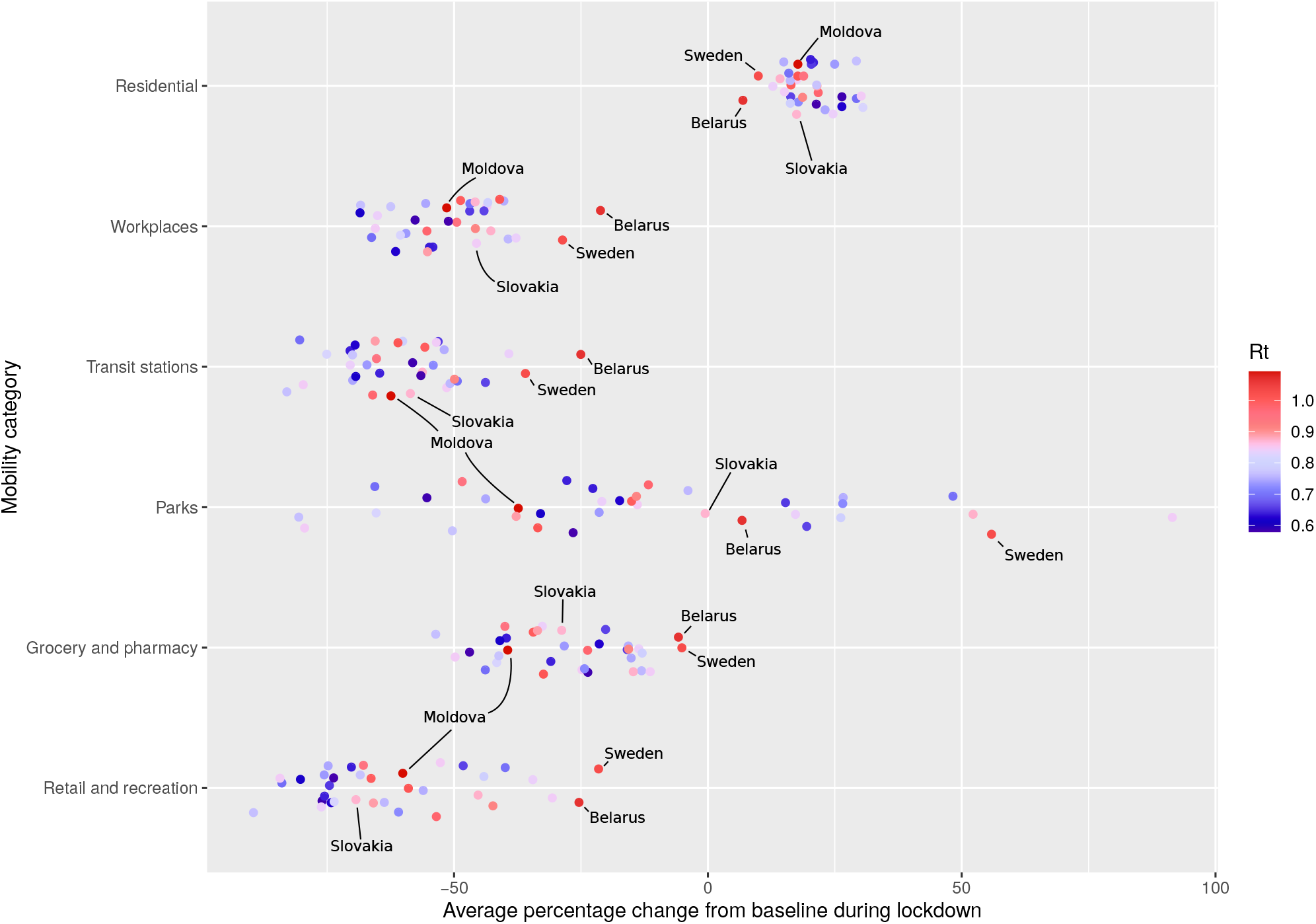
Average percentage change in mobility, compared to baseline, for six location Google mobility categories. Colour reflects basic reproduction number *R*_*t*_ during lockdown. Retail and Recreation: restaurants, cafes, shopping centers, theme parks, museums, libraries, and movie theaters; Grocery and Pharmacy: grocery markets, food warehouses, farmers markets, specialty food shops, drug stores, and pharmacies; Parks: local parks, national parks, public beaches, marinas, dog parks, plazas, and public gardens; Transit Stations: public transport hubs such as subway, bus, and train stations; Workplaces: places of work; Residential: places of residence.

By explaining only variation of *R*_*t*,2_, instead of the reduction from *R*_*t*,1_ to *R*_*t*,2_, the above model side-stepped to a large extent the colinearity that exists between changes for different mobility categories, which showed the same trend in all countries. To verify that the reduction in *R*_*t*_ as a result of the lockdown (comparing *R*_*t*,1_ before lockdown to *R*_*t*,2_ during lockdown) was also associated with mobility changes, a multivariate model that directly predicted the percentage change of *R*_*t*_ due to lockdown using average mobility changes (comparing mobility data from before *d*_1_ to after *d*_2_), was also estimated (data not shown). This model had a high explanatory power (*R*^2^ = 0.95) but as expected suffered from high colinearity. Nevertheless, in agreement with the above finding, this model also attributed an estimated 31% (95% CI 11 – 50) of the drop in *R*_*t*_ during lockdown to a reduction in mobility related to retail and recreation (*p <* 0.003).

Our findings are summarized in Figure 5 and trends were found to be robust to different assumptions of latent period duration (in the range of 2 to 4 days), and different assumptions of generation time (in the range of 4.5 to 5.9 days), see Suppl 3. Although the estimated value for *R*_*t*,2_ for the epidemic in Sweden was found to be relatively high (1.02, 95% cri 0.99 – 1.06), it was equally well explained based on mobility changes, suggesting that although not enforced, in practice the country behaved the same as other Western countries in a lockdown.

**Figure 5:**
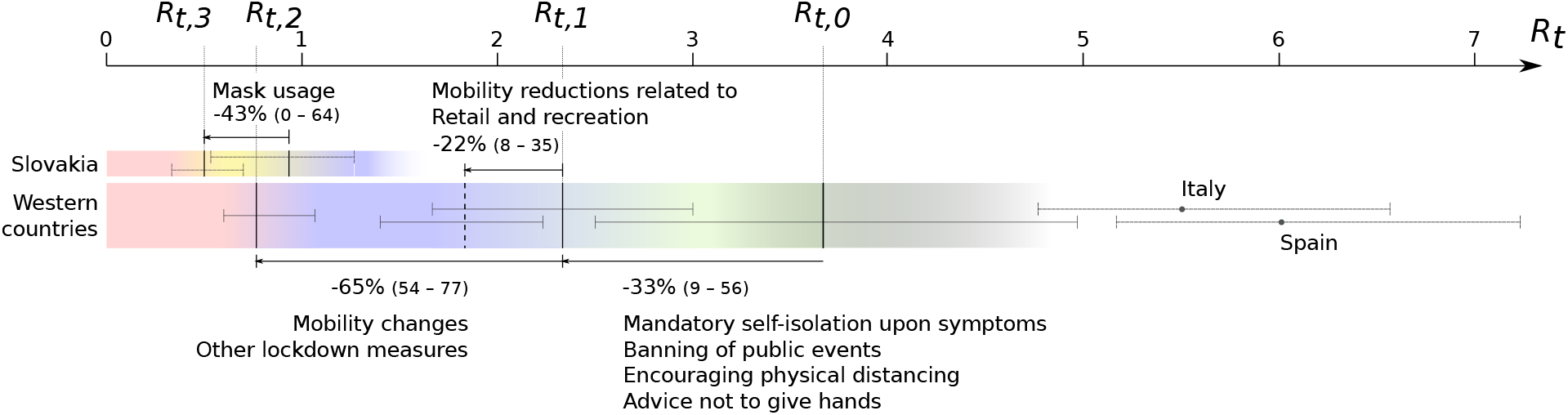
Summary of estimated contributions to the reduction of transmission in 35 Western countries. Initial basic reproduction numbers (*R*_*t*,0_), the reproduction number at start of lockdown (*R*_*t*,1_) and during lockdown (*R*_*t*,2_), and percentage reductions are shown as median values and 95% IQR. Estimates for individual countries (Italy, Spain and Slovakia) are shown as median posterior value and 95% cri.

The fact that mobility changes linked to retail and recreation were significantly associated with *R*_*t*_ both in univariate and multivariate analyses, suggests that activities related to visiting malls, bars, restaurants, or museums are linked to increased transmission and releasing those mobility restrictions should be done with care since they may carry a high risk for reigniting the epidemic. This is in line with mounting evidence of transmission being promoted by (loud) speaking or singing (bars, choir (*17*)), and longer time spent in densely populated indoor locations with low air circulation (bars, restaurants, malls, events with mass gatherings (*18*)). In the presence of general ‘physical distancing’ guidelines, which were active in all countries during lockdown, mobility changes related to workplaces (which correlated with transit stations and residential places), did not have a clear impact on *R*_*t*_, suggesting that easing of restrictions related to work may be more easily manageable. The lack of association of these other mobility parameters may suggest that rather than being associated with an individual, superspreading is mainly associated with the circumstances of the interaction. This is adding weight to the arguments that superspreading events, such as large indoor gatherings, need to be avoided.

This study has several limitations. The use of a compartment model in conjunction with a piecewise linear model for *R*_*t*_ is necessarily an approximation, and more complicated temporal changes in transmission may have occurred in reality, but still it served the purpose of examining the impact of different measures over time. Epidemiological parameter estimates may have been impacted by shifts in age distribution of cases over time, given that COVID-19 has a distinct age risk profile, and also by variation in data accuracy between countries. By limiting the study to Western countries which coincided more or less in time, differences related to culture, climate, and google mobility data interpretation, may have been less of a confounding factor.

## 1 Methods

The following equations describe the dynamics of individuals in each of the four compartments of a standard SEIR model (see Figure 6):

**Figure 6:**
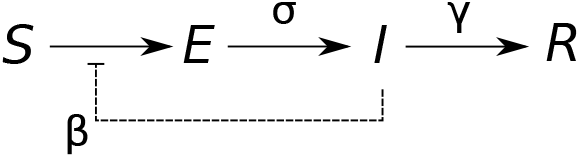
Structure of a standard SEIR compartment model with four compartments: susceptible (*S*), exposed (*E*), infectious (*I*), and removed (recovered or deceased, *R*). Susceptible individuals become latently infected by infectious individuals, with transmission rate *β*. Latently infected become infectious at rate *σ*. Infectious people are removed at rate *γ*.

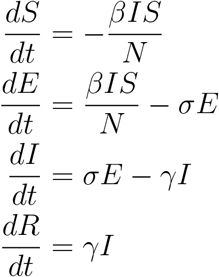

The differential equations of the SEIR compartment models were numerically integrated using deSolve (*19*). The duration of the latent period *T*_lat_ = 1*/σ* was fixed to 3 days, and the infectious period *T*_inf_ = 1*/γ* to 5.2 days based on literature (*20, 21*) (robustness analyses for other parameter values were conducted, see Suppl 3).

The dates *d*_1_ and *d*_2_ which mark the transition period for mobility changes were estimated from mobility data reports (*22*), by fitting a step function with a linear transition period through the sum of mobility changes related to transit stations and workplaces (see Suppl 1). This was motivated by the adoption of teleworking as a measure in most countries, even in those (like Sweden) which had the lightest lockdown regimen. The value for *β* was modeled as a piecewise linear function of time (Figure 1).

We wanted a model that does not require assumptions on timing or number of introductions per country. Instead, the models were seeded with an initial single exposed individual and it was assumed that the estimated date *d*_0_, marking the first change in value of *β*, was linked to a co-estimated threshold for cumulative deaths. This method had the benefit of avoiding any bias on estimated *R*_0_ values because of assumptions on introduction time and numbers, while being more efficient to sample than time of first infection.

Suppl 3 Table 1 lists all parameters used in the model and during the estimation from data, with their values (either a constant, or a prior distribution for parameters that were estimated).

To estimate the model parameters, incidence data of diagnosed cases and deaths was used (ECDC, https://opendata.ecdc.europa.eu/covid19/casedistribution/csv, accessed on 6 June), within a Bayesian framework. The daily incidence of diagnosed cases and deaths was evaluated against the prediction based on the simulation of the SEIR model, using a negative binomial distribution. The posterior distributions of model parameters were estimated in R using MCMC with Metropolis coupling (20 chains at different temperatures) (*23–26*).

Univariate and multivariate linear models were used for quantifying the effect of the average values of Google Mobility data (*22*) over the time period prior to lockdown to *R*_*t*,1_ and during lockdown to *R*_*t*,2_. Colinearity was assessed by calculating covariance-inflation factors (*27*). Statistical analysis was done in RStudio (*28*).

All data files, R scripts and analysis steps are described in Suppl 2.

## Data Availability

All data and analysis software are openly available.

https://github.com/kdeforche/epi-western-analysis

## 2 Acknowledgments

The resources and services used in this work were provided by the VSC (Flemish Supercomputer Center), funded by the Research Foundation - Flanders (FWO) and the Flemish Government. VM was supported by the ELTE Institutional Excellence Program (NKFIH-1157-8/2019-DT) and a COVID-19 grant (2020-2.1.1-ED-00001) of the National Research, Development and Innovation Office.

## 3 Author contributions

K.D. contributed to the Bayesian analyses and conception of the study; J.V. contributed to the statistical analyses; V.M. contributed to the Bayesian analyses and epidemiological interpration; A.M.V. contributed to the virological and epidemiological interpretation and revisions of the manuscript.

